# Clinical Outcomes Following Optical Coherence Tomographic versus Intravascular Ultrasound-Guided Percutaneous Coronary Intervention: A Meta-Analysis of Randomized Controlled Trials

**DOI:** 10.1101/2024.04.18.24306053

**Authors:** Sheriff N. Dodoo, Sammudeen Ibrahim, Abdul-Fatawu Osman, Rayan Salih, Vikas Kilaru, Dennis Kwaku Ampadu, Afia S. Dodoo, Ugochukwu Egolum, Nima Ghasemzadeh, Ronnie Ramadan, Gregory Giugliano, Glen Henry, Uzoma Ibebuogu, Habib Samady

## Abstract

**Background:** Optical coherence tomography (OCT) and intravascular ultrasound (IVUS) are adjunctive intracoronary imaging modalities used for optimizing the implantation of coronary stents. However, the impact of the choice of OCT versus IVUS on clinical outcomes and periprocedural complications is unclear.

**Objective:** To perform a meta-analysis of all vetted randomized controlled trials comparing OCT-guided versus IVUS-guided percutaneous coronary intervention.

**Methods:** We queried MEDLINE, Cochrane Library, Scopus, and clinicalTrials.gov databases from their commencement to February 2024 for all randomized controlled trials that compared OCT-guided versus IVUS-guided percutaneous coronary interventions. The primary endpoint was major adverse periprocedural events (MAPE), a composite of stent thrombosis (ST), distal embolization (DE), and distal edge dissection (DED). The secondary endpoints included stent thrombosis, distal embolization, distal edge dissection, Major-adverse-cardiac events (MACE)-[a composite of cardiac death, Target vessel myocardial infarction (TVMI), and target vessel revascularization (TVR)], all-cause mortality, cardiac death, TVMI, TVR, and nonfatal stroke. The odds ratio (OR) with a 95% confidence interval (CI) was analyzed using a random-effect model.

**Results:** Seven randomized controlled trials were included in the analysis, and 4446 patients were enrolled. OCT was associated with lower MAPE (OR: 0.65, CI:0.47-0.91, p= 0.01) compared to IVUS. ST, DE, and DED were similar between OCT and IVUS. There were no significant differences in MACE (OR: 0.86, CI:0.64-1.16, p= 0.32), all-cause mortality (OR: 0.83, CI:0.42-1.66, p= 0.60), Cardiac death (OR: 0.62, CI:0.20-1.89, p= 0.40), TVMI (OR: 0.69, CI:0.33-1.46, p= 0.33), TVR, (OR: 1.09, CI:0.70-1.71, p= 0.70), and non-fatal stroke (OR: 1.82, CI:0.67-4.95, p= 0.24) one year following the index procedure.

**Conclusion:** Optical coherence tomographic-guided PCI was associated with lower major adverse peri-procedural events (MAPE), including stent thrombosis, distal embolization, and distal edge dissection, compared to intravascular ultrasound-guided PCI. However, there was no difference in overall major adverse cardiac events, target vessel myocardial infarction, target vessel revascularization, and nonfatal stroke.

## 1. INTRODUCTION

Optical coherence tomography (OCT) is an adjunctive intracoronary imaging modality used to define the morphology of coronary plaque, guide the placement of coronary stents, and optimize percutaneous coronary intervention (PCI) [1]. Intravascular ultrasound (IVUS) is a valuable alternative imaging technique for PCI guidance. OCT has superior resolution compared to IVUS, allowing more precise plaque characterization and diagnosis of periprocedural complications, including malapposition and distal edge dissection [1,2]. However, the improved resolution of OCT is at the expense of decreased imaging depth [2].

Multiple clinical trials have demonstrated superior clinical benefit with IVUS-guided PCI compared with angiographically guided PCI [3-5]. The Implantation of larger caliber stents, prompt diagnosis and treatment of gross malapposition, and distal edge dissection in the IVUS cohorts are some of the proposed mechanisms underlying the improved clinical outcomes observed compared to the angiographic cohorts. Consequently, IVUS has found clinical utility in high-risk PCI, particularly in left main lesions [6].

Recently, Kang and his colleagues reported findings of a prospective open-label pragmatic trial comparing OCT-guided PCI with IVUS-guided PCI, which showed OCT was non-inferior to IVUS for the incidence of a composite of death from cardiac causes, target vessel-related myocardial infarction or ischemia-driven target vessel revascularization at one year [7]. Ali and colleagues also showed that OCT-guided PCI significantly improved the acute minimal stent area, compared with angiography-guided PCI among patients undergoing coronary revascularization. However, this did not improve clinical outcomes in a prospective, single-masked, randomized controlled trial (RCT) comparing the efficacy and safety of OCT-guided PCI with angiography-guided PCI [8].

The periprocedural benefits of OCT-guided PCI compared to IVUS-guided PCI are limited. We sought to perform a meta-analysis of randomized controlled trials comparing the effectiveness and safety of OCT-guided PCI with IVUS-guided PCI.

## 2. METHODS

This meta-analysis complied with the preferred reporting items for systematic reviews and meta-analyses (PRISMA) and the Cochrane protocol [9,10]. This study was exempted from institutional board review board approval as the included RCTs were all publicly available, and their clinical data were de-identified. Additionally, the study was registered under the International Prospective Register of Systematic Reviews (PROSPERO) with registration number CRD42024528072.

### 2.1 Data sources and query strategy

MEDLINE, Cochrane Library, Scopus, and clinicalTrials.gov databases were queried for all RCTs published through February 2024, comparing the effectiveness and safety of OCT-guided PCI versus IVUS-guided PCI. The following keywords were used for the search: “Optical coherence tomography,” “OCT coronary,” “Intravascular ultrasound,” “IVUS coronary,” “IVUS-guided PCI,” “OCT-guided PCI,” “Intravascular ultrasound guided-PCI” and “Optical coherence tomography guided-PCI.”

### 2.2 Study selection and endpoints

Two authors (SND and SI) screened all the studies independently and in tandem. Seven RCTs that satisfied the inclusion criteria were included in the final analysis. The inclusion criteria included (a) only RCT, (b) studies of patients 18 years and above, (c) study compared OCT with IVUS, including three-arm RCT that compared OCT versus IVUS versus angiography in which the three comparative groups were clearly defined, and delineated and (e) RCT reported at least one of the endpoints of interest: major adverse periprocedural events (MAPE), Stent thrombosis (ST), distal embolization (DE), distal edge dissection (DED), major adverse cardiac event (MACE), all-cause mortality, cardiac death, target vessel MI (TVMI), target vessel revascularization (TVR), and nonfatal stroke. The primary endpoint of interest was MAPE, a composite of ST, DE, and DED. The secondary endpoints were ST, DE, DED, MACE-a composite of cardiac death, TVMI and TVR, all-cause mortality, cardiac death, TVMI, TVR, and nonfatal stroke. The periprocedural events were defined as outcomes reported during the initial hospitalization or within 30 days of the index procedure. The long-term outcomes were adjudicated as adverse events occurring from >30 days of the index procedure and up to 1 year of follow-up.

### 2.3 Data extraction and evaluation of study quality

After all duplicated data were identified and removed, all the relevant studies were exported to the Endnote Reference Manager (version x5: Clarivate Analytics). The prespecified variables from the selected RCTs were incorporated into a dataset independently and in tandem by the first two authors (SND and SI). The incorporated extraction was independently verified by a third author (UE). The clinical variables exported included baseline characteristics when available and data on the endpoints of interest, including MAPE, ST, DE, DED, MACE, all-cause mortality, cardiac death, TVMI, TVR, and nonfatal stroke. The quality of the included RCTs was assessed using the modified Cochrane Collaboration’s risk of bias tool. The publication bias was also assessed using a visual inspection of the funnel plot.

### 2.4 Statistical analysis

All the statistical analyses were done using Review Manager (version 5.4.1; Copenhagen: The Nordic Cochrane Center, the Cochrane Collaboration, 2014). Mantel-Haenszel Odds Ratio (OR) was used to assess categorical variables estimated with a 95% confidence interval (CI). Heterogeneity between study protocols was assessed using Cochran’s Q and I^2^ statistics. Heterogeneity was adjudicated with a P value <0.05 and an I2 value of >50%. The pooled effect estimate was derived using a random effects model for study endpoints with inverse variance weights. Fixed-effect model was used for data analysis if I^2^ statistics <50%; otherwise, the random-effect model was used. The included RCTs in the analyses reported 1-year outcome data, while the periprocedural events were provided from the initial hospitalization up to 30 days from the index procedure. The forest plots were provided for both periprocedural events and the 1-year outcomes of interest.

## 3. RESULTS

### 3.1 characteristics of study participants

The study flow diagram for selecting the RCTs included in the final analysis is shown in Figure 1. Seven RCTs involving 4446 patients were selected for the final analysis, with 1977 in the OCT arm and 2469 in the IVUS arm [3, 11-16]. All the seven RCTs were published from 2012 to 2023. The baseline characteristics of the participants of included RCTs are summarized in the supporting information Table S1. The included RCTs showed minimal publication bias, as depicted by the symmetrical funnel plot shown in supporting information Figure S1. All the included studies were of sufficient quality.

**Figure 1:**
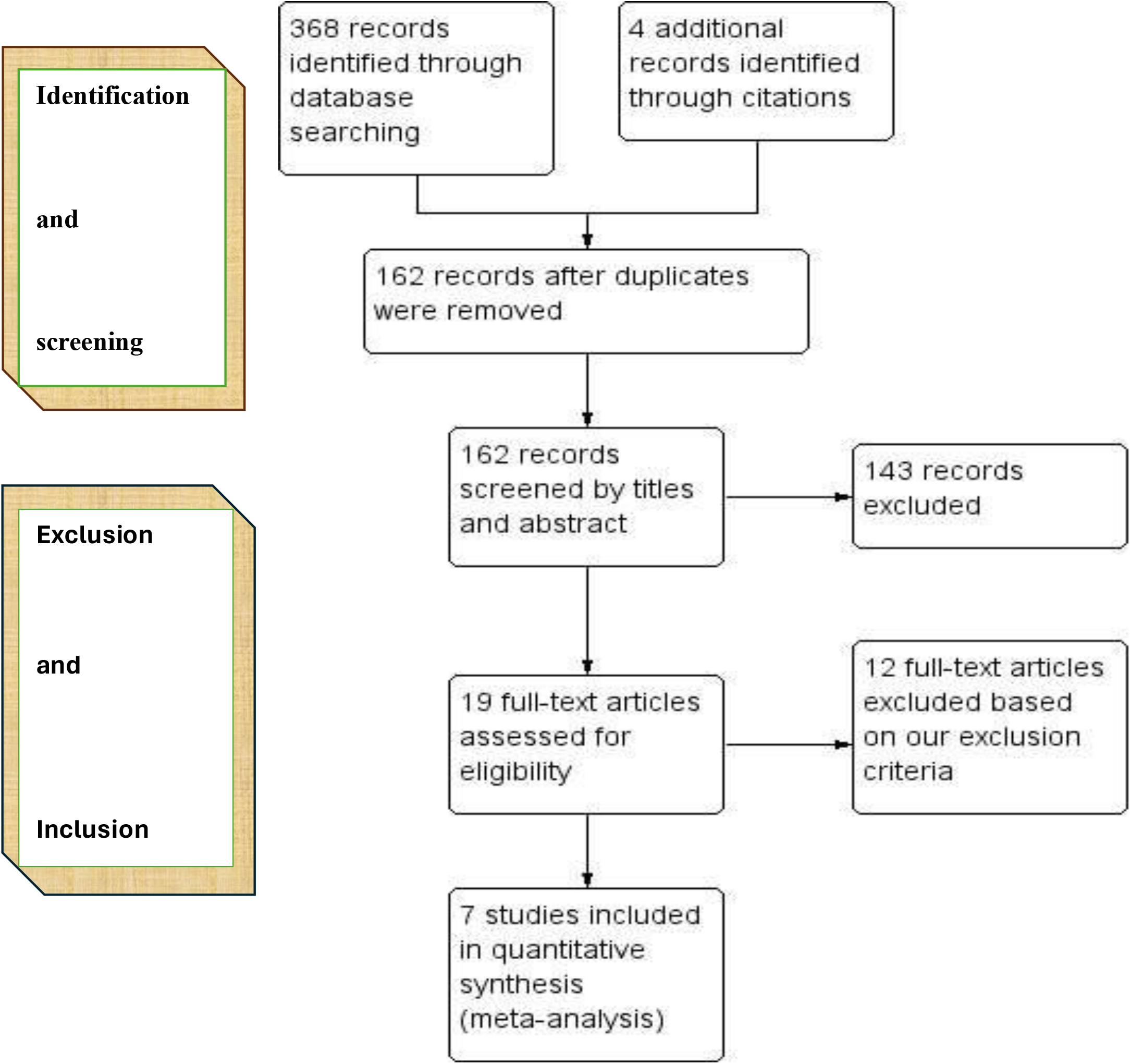
Preferred Reporting Items for Systematic Reviews and Meta-Analyses (PRISMA) study flow diagram

### 3.2 Results of meta-analysis

The primary endpoint of interest was a major adverse periprocedural event (MAPE), a composite of ST, DE, and DED. Six of the seven included RCTs reported ST, DE, and DED, allowing for composite MAPE analysis. The results of the meta-analysis are shown in Figure 2. OCT was associated with lower MAPE when compared to IVUS (OR: 0.65, CI:0.47-0.91, p= 0.01). OCT was associated with a trend toward a lower ST than IVUS (OR:0.44, CI:0.19-1.03, p=0.06). There was no significant difference in DE (OR:0.32, CI:0.06-1.60, p=0.16) and DED (OR:0.92, CI:0.56-1.50, p=0.73) when OCT was compared to IVUS.

**Figure 2:**
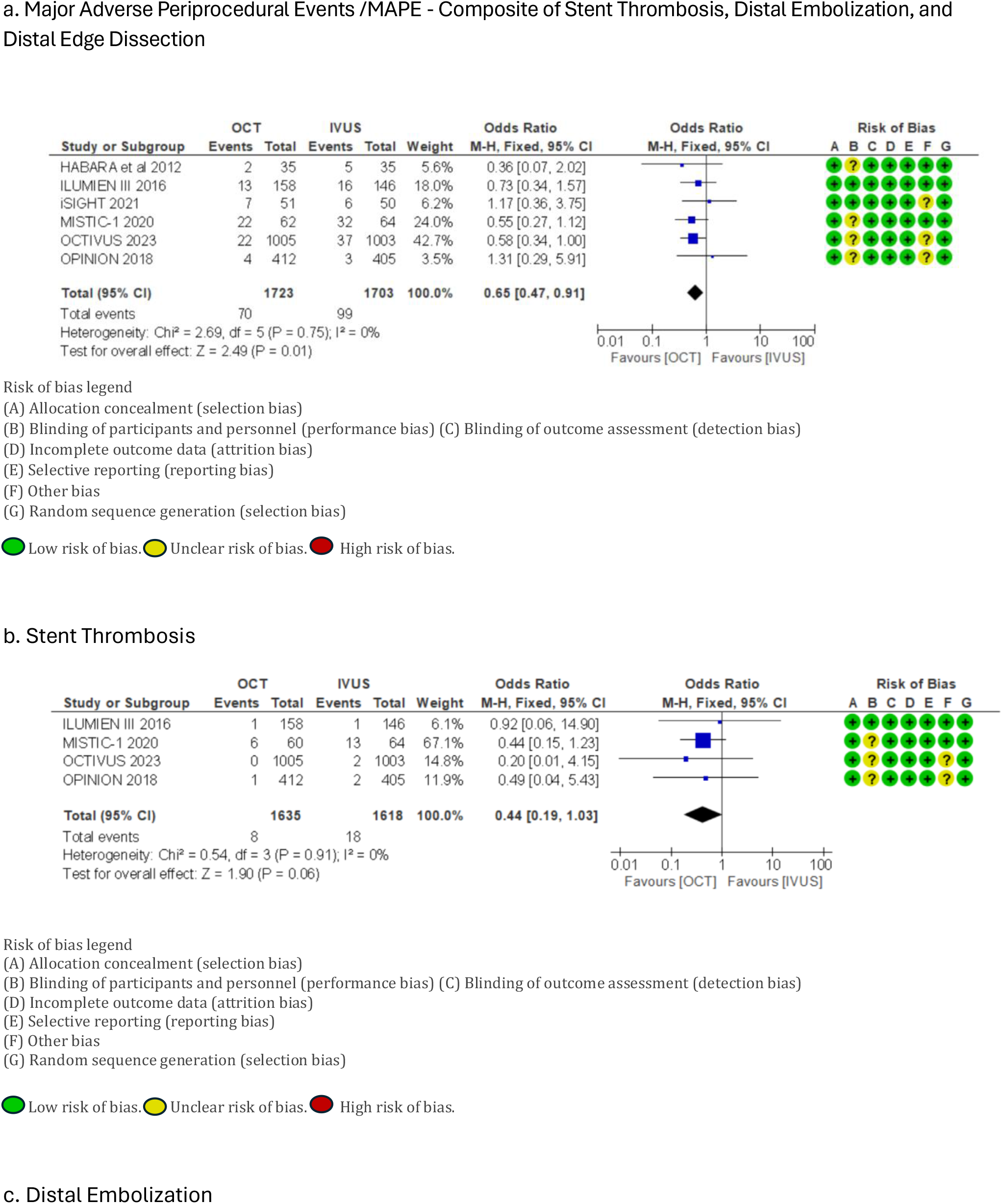
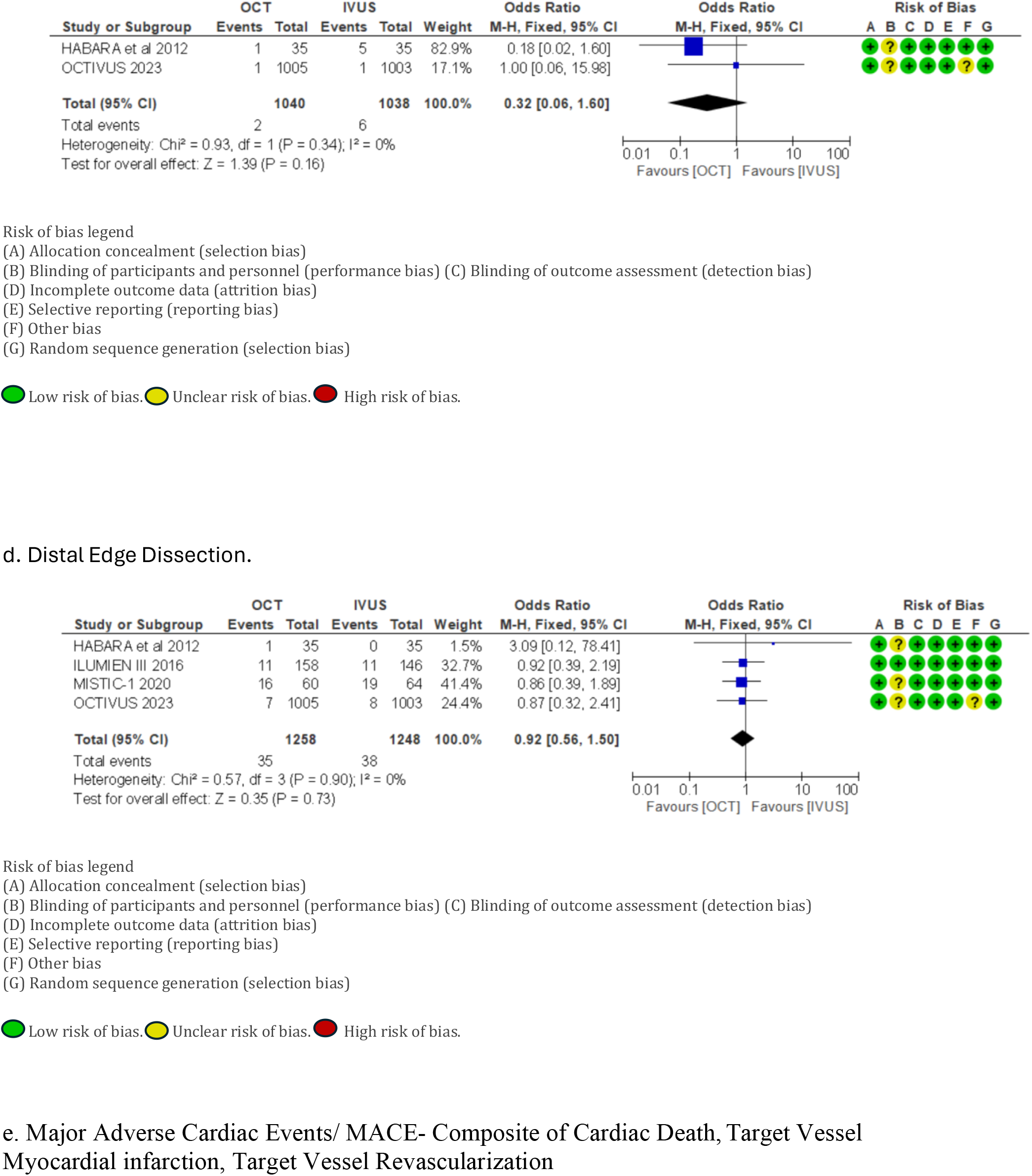
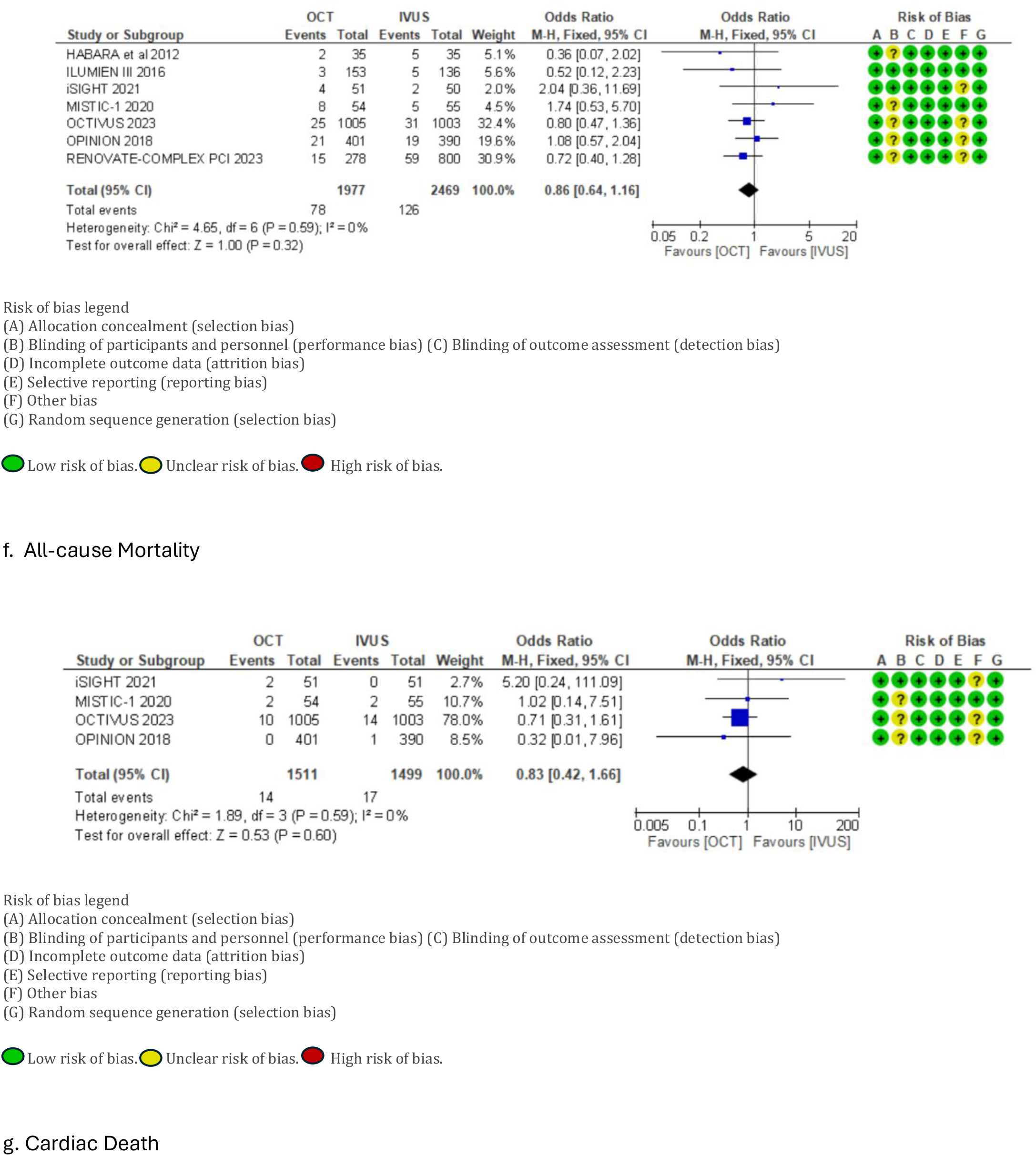
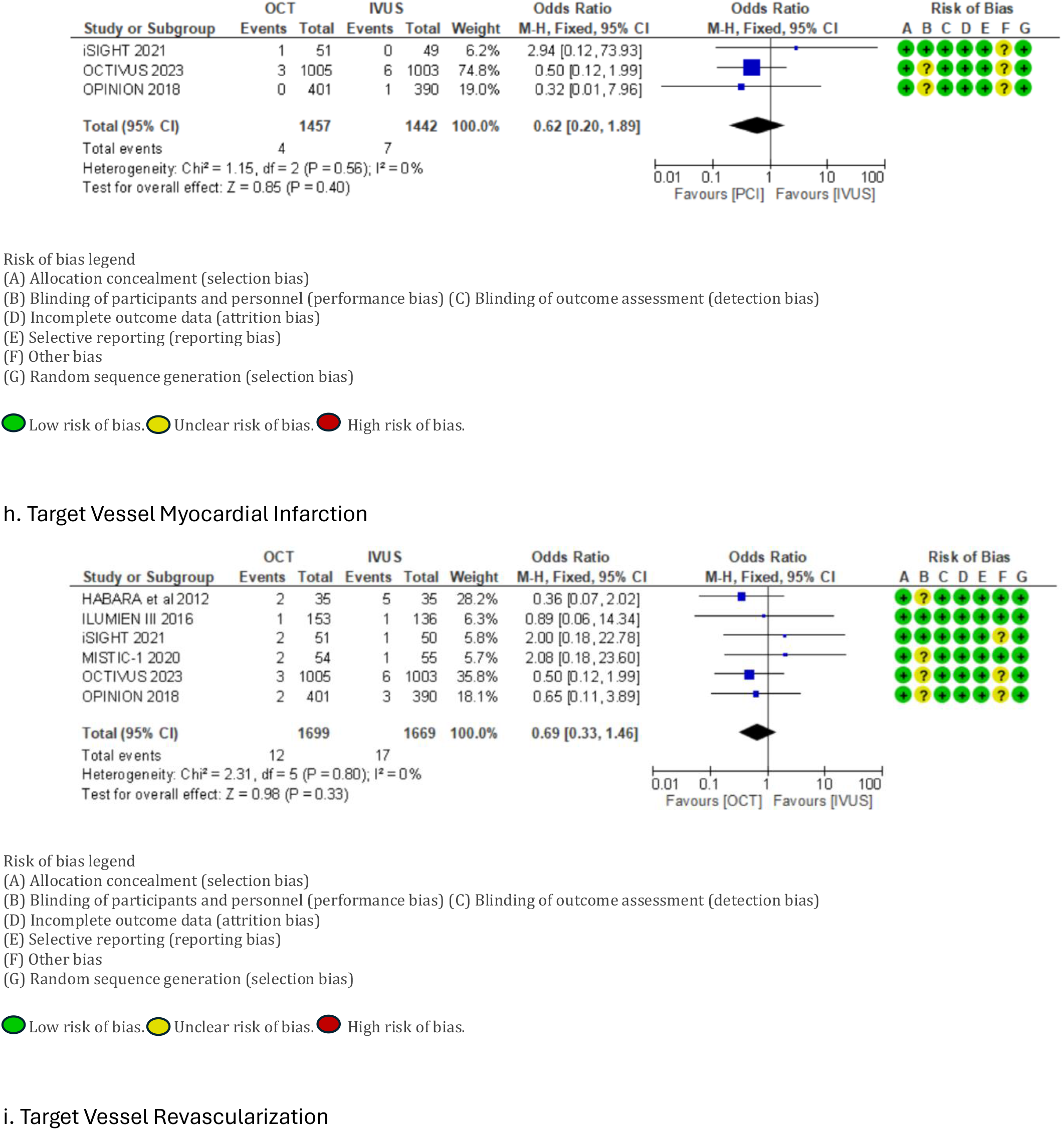
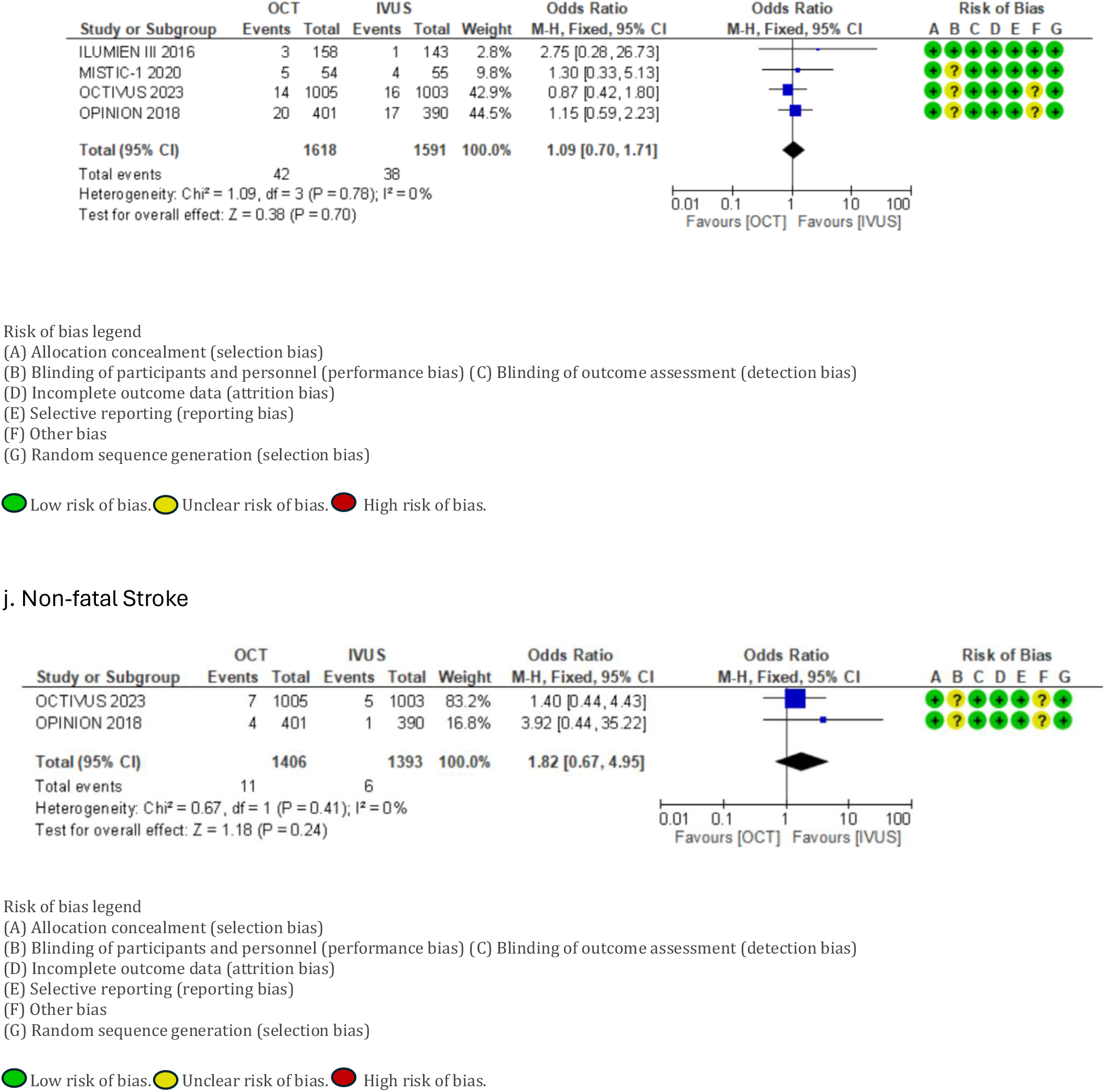
Forest plot of the pooled analysis comparing optical coherence tomography and intravascular ultrasound following percutaneous coronary interventions (a.) Major Adverse Periprocedural Events/MAPE-Composite of Stent Thrombosis, Distal Embolization, and Distal Edge Dissection (b.) Stent Thrombosis (c.) Distal Embolization (d.) Distal Edge Dissection (e.) Major Adverse Cardiac Events/MACE-Composite of Cardiac Death, Target Vessel Myocardial Infarction, Target Vessel Revascularization (f.) All-cause Mortality (g.) Cardiac Death (h.) Target Vessel Myocardial Infarction (i.) Target Vessel Revascularization (j.) Non-fatal Stroke

There was also no difference in MACE (OR:0.86, CI:0.64-1.16, p=0.32), All-cause mortality (OR:0.83, CI:0.42-1.66, p=0.60), cardiac death (OR:0.62, CI:0.20-1.89, p=0.40), TVMI (OR:0.69, CI:0.33-1.46, p=0.33), TVR (OR:1.09, CI:0.70-1.71, p=0.70), and non-fatal stroke (OR:1.82, CI:0.67-4.95, p=0.24) when OCT was compared to IVUS.

## 4. DISCUSSION

In this present meta-analysis of RCTs comparing OCT-guided versus IVUS-guided PCI, we report several significant findings: (1) OCT-guided PCI was associated with lower MAPE compared with IVUS-guided PCI; (2) In the analysis of seven RCTs, OCT-guided PCI showed a trend for lower stent thrombosis when compared with IVUS-guided PCI; (3) No difference in DE, DED, MACE, All-cause mortality, cardiac death, TVMI, TVR, and nonfatal stroke was detected when OCT-guided PCI was compared to IVUS-guided PCI.

Stent thrombosis, distal embolization, and distal edge dissection are major complications following PCI. Stent thrombosis, a potentially life-threatening and fatal outcome following PCI [17], accounts for a mortality rate between 5 and 45%, as well as a recurrence rate of 15–20% at five years [18,19,20]. Distal embolization occurs in up to 15% of patients undergoing primary PCI. It has been associated with poor reperfusion, larger infarct size, lower left ventricular ejection fraction, and an unfavorable five-year survival [21]. Depending on the coronary imaging modality used, the incidence of distal edge dissection following coronary stent implantation ranges from 7.8–19.0% with IVUS [22,23] and 19.0% to 39.1% with OCT [24,25,26]. Distal edge dissection detected by IVUS following PCI has been associated with worse clinical outcomes, including ST and target lesion revascularization [28]. Distal edge dissection diagnosed by OCT has also been associated with MACE [29].

Intracoronary imaging in guiding stent implantation includes avoiding periprocedural complications, including ST, DE, and DED, and where they occur, allowing for prompt recognition and management [30]

In the Ilumien III trial, Ali and colleagues found that PCI with OCT guidance resulted in fewer untreated major dissections than the IVUS group and fewer areas of major stent malapposition than IVUS guidance. Similarly, untreated major stent malapposition after PCI was more frequent with IVUS and angiography guidance than with OCT guidance. However, the OCT-detected plaque or thrombus protrusion frequency was not significantly different between groups [12]. In the OCTIVUS trial [15] comparing OCT-guided vs. IVUS-guided PCI, the incidence of major procedural complications requiring active intervention was lower in the OCT group than in the IVUS group. The authors attributed this difference in procedural complication to a more aggressive interventional approach in the IVUS arm. The lumen-based approach commonly used for OCT-guided PCI and a vessel-based (using external elastic lamina) strategy usually used for IVUS-guided PCI may have accounted for the differences in procedural complications, including distal edge dissection and distal embolization. However, the IVUS group had a larger maximal stent area than the OCT group.

These findings were in keeping with the findings of this present meta-analysis, which showed that the odds of major adverse periprocedural event (MAPE), a composite of ST, DE, and DED, were lower in the OCT arm compared to the IVUS arm.

The higher OCT resolution than IVUS imaging confers greater sensitivity for detecting postprocedural dissections, malapposition, and thrombus. It also allows for better evaluation and assessment of coronary plaque morphology, including calcified lesions. IVUS allows more complete vessel and plaque visualization at the expense of a lower resolution. Additionally, the pullback speed of OCT is faster than that of IVUS, resulting in quicker and more efficient co-registration with automated measurements of luminal and lesion dimensions with OCT. This may facilitate a rapid comprehensive evaluation of a long segment of stented vessels; hence, OCT guidance was associated with a shorter PCI time [15].

Historically, a significant limitation of OCT is the use of contrast to clear blood from the intracoronary lumen, facilitating the penetration of infrared light to image the bloodless luminal bed. The suggestion was that this would translate into a higher incidence of contrast-induced nephropathy in cases that used OCT intracoronary imaging compared with IVUS. However, in the OCTIVUS trial, although, the amount of contrast used during the procedures was higher in the OCT group than in the IVUS group, it did not increase the incidence of contrast-induced nephropathy in the OCT group.

Data from this meta-analysis also showed no difference in DE, DED, MACE, all-cause mortality, cardiac death, TVMI, TVR, and nonfatal stroke detected when OCT-guided PCI was compared to IVUS-guided PCI one year following the index procedure. This finding agreed with prior studies, which showed similar outcomes, including the ILUMIEN III, iSIGHT, MISTIC-1, OCTIVUS, OPINION, and RENOVATE-COMPLEX PCI trials.

A prospective multicenter RCT of OCT-Guided Coronary Stent Implantation Compared with Angiography (ILUMIEN IV: OPTIMAL PCI) was conducted to determine whether OCT-guided PCI would improve procedural and clinical outcomes as compared with angiography-guided PCI. This pivotal trial showed that OCT guidance resulted in a larger minimum stent area after PCI than angiography guidance. However, there was no apparent difference in a composite of death from cardiac causes, target-vessel myocardial infarction, or ischemia-driven target-vessel revascularization at two years [31]. There was also few OCT imaging–and procedure-related complications, and stent thrombosis through 2 years appeared to be less frequent with OCT guidance than with angiography guidance.

Interpretation of the result of this meta-analysis should be done within the confines of several limitations. First, this meta-analysis was done assuming that the baseline characteristics of the participants of the seven RCTs included were homogenous and baseline. Additionally, the patient-level data of the RENOVATE-COMPLEX PCI was not available. While this heterogeneity in baseline characteristics of the study participants could have confounded the analysis results, there was a relatively low statistical heterogeneity in comparative groups. Second, there were differences in the study protocol when applying OCT and IVUS to guide the placement of stents.

In contrast, some studies used the average of proximal and distal luminal dimensions as a reference for sizing the stents placed; others used the distal luminal dimension. Effects of the differences in the study protocols of the individual included RCTs were not assessed but could affect the results of this meta-analysis. The random effect model was used to mitigate the effects of the differences in study protocol on the results of this meta-analysis. Fourth, there was no available data on the experience and expertise of the operators, which may affect rates of peri-operative events in each arm. Finally, six of the seven included RCTs reported stent thrombosis, distal embolization, and distal edge dissection, allowing the composite analysis of ST, DE, and DED, as denoted as the MAPE, the primary endpoint of the meta-analysis.

## 5. CONCLUSION

This meta-analysis showed that OCT-guided PCI was associated with lower MAPE, including ST, distal embolization, and distal edge dissection, compared to IVUS-guided PCI. However, there was no difference in overall MACE, TVMI, TVR, and nonfatal stroke. While IVUS is a safe and effective adjuvant to guiding the optimal placement of coronary stents, OCT is a reasonable option. These findings should be considered hypothesis-generating as more high-fidelity trials are needed to elucidate the mechanism underlining these outcomes.

## Data Availability

The data that supports the findings of this study are available in the supplementary material of this article.

